# Prevalences and healthcare expenditures related to 58 health conditions from 2012 to 2017 in France: diseases and healthcare expenditure mapping, a national population-based study

**DOI:** 10.1101/2020.09.21.20198853

**Authors:** Antoine Rachas, Christelle Gastaldi-Ménager, Pierre Denis, Thomas Lesuffleur, Muriel Nicolas, Laurence Pestel, Corinne Mette, Jérôme Drouin, Sébastien Rivière, Ayden Tajahmady, Claude Gissot, Anne Fagot-Campagna

## Abstract

**Background:** Description of the prevalence of diseases and resources mobilized for the management of each disease is essential to identify public health priorities. We described the prevalences of 58 health conditions and all reimbursed healthcare expenditure by health condition in France between 2012 and 2017.

**Methods and Findings:** All national health insurance general scheme beneficiaries (87% of the French population) with at least one reimbursed healthcare expenditure were included from the French national health database. We identified health conditions (diseases, episodes of care, chronic treatments) by algorithms using ICD-10 codes for long-term diseases or hospitalisations, medications or medical procedures. We reported crude and age and sex-standardized annual prevalences between 2012 and 2017, and total and mean (per patient) reimbursed expenditure attributed to each condition without double counting, and according to the type of expenditure.

In 2017, in a population of 57.6 million people (54% of women, median age: 40 years), the most prevalent diseases were diabetes (standardized prevalence: 5.8%), chronic respiratory diseases (5.5%) and chronic coronary heart disease (2.9%). Diseases concentrating the highest expenditures were active cancers (10% of total expenditure of €140.1 billion), mental illness (10%; neurotic and mood disorders: 4%; psychotic disorders: 3%), and chronic cardiovascular diseases (8%). Between 2012 and 2017, the most marked increase in total expenditure concerned liver and pancreatic diseases (+9.3%), related to the increased drug expenditure in 2014. Conversely, the increase in the number of patients (and the prevalence) explained the majority of the increase of total expenditures for cardiovascular disease, diabetes and mental illness.

**Conclusions:** These results showed a regular increase of the prevalence and expenditure of certain chronic diseases, probably related to ageing of the population, and increased expenditures related to major therapeutic innovations. The Diseases and Healthcare Expenditure Mapping therefore enlightens decision-makers in charge of public health and health accounts.

## Introduction

In high-income countries, increasing life expectancy and changing lifestyles have led to an increased prevalence of chronic diseases and the number of years lived with disease. (1) To address the many challenges resulting from longer life expectancy (medical, related to healthcare organization, financial) and global health crisis such as COVID-19 pandemic, we need to reinforce the resilience of health systems, defined by the OECD as the capacity of health systems to adapt efficiently to changing economic, technological and demographic environments. (2) In a context of increasing financial constraints, a health system must be able to meet the dual human and economic challenge to ensure its own sustainability. Description of the prevalence of diseases and resources specifically mobilized for the management of each disease is essential to identify public health priorities and to more clearly understand some of the mechanisms underlying healthcare expenditure. (3–5)

Over the last decade, tools have been developed to describe morbidity, prevalence of diseases and/or healthcare expenditure related to these diseases. The Global Burden of Disease (GBD) provides, for each country of the world, estimations of incidences, prevalences, numbers of disability-adjusted life years (DALYs), years of life lost (YLLs), and years lost due to disability (YLDs) and describes the causes of death. (1,6) The Health Expenditures by Diseases and Conditions project is designed to describe expenditure by disease in 14 European countries. (7) At the national level, claims data used for healthcare billing are increasingly used to describe expenditure by disease (4,5,8–12) and morbidity (13,14) and sometimes contribute to GBD data. However, the lack of individual information sometimes requires the use of modelling, especially to estimate prevalences or YLDs (as in the GBD) or to group diseases into large categories. Another common limitation is the lack of representativity of the study population compared to the general population when health insurance is not universal. Finally, only very few studies have simultaneously estimated the prevalences of diseases and described the expenditure related to each disease (15).

In France, the National Health Data System (“*Système National des Données de Santé, SNDS*”) contains extensive individual information (including inpatient, outpatient, diagnosis and medication data) for almost the entire French population, as public health insurance coverage is mandatory. (16) For these reasons, SNDS has been increasingly used for clinical and health services research over recent years. (17,18) Since 2012, SNDS has been used to develop a standardized tool, the Diseases and Health Expenditures Mapping (DHEM), the results of which are published in an annual report to parliament, which is used to prepare the Social Security Funding Act. The main objective of the DHEM was to describe nationwide, annual expenditures by health condition for a broad spectrum of health conditions (58 treated diseases, chronic treatments and episodes of care) with no double counting, previously described as a general cost-of-illness study, a conceptually relevant method for a political objective. (19) The secondary objective was to estimate the prevalences of these health conditions in healthcare users, globally and by age and sex. In this paper, we present, for the first time, the methodology of the DHEM, the main results for 2017 and their variations between 2012 and 2017.

## Methods

### Data source: the French National Health Data System (SNDS)

French national health insurance provides mandatory health insurance coverage by means of several schemes, depending on occupational class, that cover the entire population. Individual data from administrative forms and reimbursement claims have been prospectively recorded in a common data warehouse, SNDS, since 2003. (16) SNDS contains demographic data, including vital status, and exhaustive information on pharmacy claims and the type of outpatient services or procedures reimbursed (without their results). In outpatient settings, physician-reported diagnoses are available only for beneficiaries with “*Affection de Longue Durée*” status, which waives copayments related to the treatment of specific long-term diseases (LTD). Each LTD diagnosis is validated individually by a national health insurance physician according to regulations. SNDS also includes the French Hospital Discharge Database (“*Programme de Médicalisation des Systèmes d’Information*”), containing inpatient diagnoses and procedures, from 2006 to 2017 at the time of this study. Overall, information from pharmacy claims and the use of both inpatient and outpatient healthcare services, including related expenditure, is available. Diagnoses (LTD and hospital discharge) are recorded according to the ICD 10th Revision (ICD-10) codes and drugs are recorded according to the Anatomical Therapeutic Chemical (ATC) index.

All SNDS data are anonymous and individually linkable. Access to data is subject to prior training and authorisation and needs approval by the independent French data protection authority (“*Commission Nationale Informatique et Libertés*”). Our public institution has permanent access to these data in application of the provisions of articles R. 1461-12 et seq. of the French Public Health Code, therefore ethical board approval was not required.

### Study design

The DHEM is equivalent to an annually repeated cross-sectional population-based study, without sampling since all eligible individuals are included. As the method is progressively modified each year, the study is repeated on all years studied at each new version. As the linkage of each individual over time in the SNDS was substantially improved in 2012, the present study was therefore conducted over the years 2012 to 2017.

### Study population

The DHEM has included all people with at least one reimbursement for healthcare delivery during the year studied. The DHEM has been restricted to beneficiaries of the general scheme or other specific health insurance schemes (“*Sections Locales Mutualistes, SLM”*), for whom LTD status was comprehensively recorded during the study period. These schemes cover about 57 million people (87% of the French population).

### Health conditions

Algorithms have been developed in order to identify 58 health conditions (grouped in 15 categories) from the medical information available in the SNDS (summarised in a previous article (20)). As the primary objective of the DHEM was to attribute reimbursed expenditures between various health conditions, the algorithms were not designed to estimate the incidence and prevalence of diseases (including people with a disease that is either not diagnosed or not treated), but to identify populations with treated diseases, chronic treatments and frequent, serious or expensive care episodes. All algorithms were submitted to expert review and have been subsequently adapted. Continuing improvement of the DHEM algorithms has also benefited from the work conducted by REDSIAM, a network of SNDS expert users, which records and compares existing algorithms. (21) Detailed definitions and expert review of the DHEM algorithms are publicly available in French (https://www.ameli.fr/l-assurance-maladie/statistiques-et-publications/etudes-en-sante-publique/cartographie-des-pathologies-et-des-depenses/methode.php).

These algorithms were based on the following elements: LTD ICD-10 code; ICD-10 code of diagnoses related to hospitalisations during the year studied (or during up to 5 previous years, depending on the algorithm); drugs that are specific to certain chronic diseases (diabetes, HIV infection, Parkinson’s disease, etc.); and, for several diseases, laboratory tests (HIV infection), medical procedures, lump sums or diagnosis-related group (end-stage renal disease). Chronic treatments were defined by the presence of at least three drug dispensings during the year (six dispensings for “analgesics or non steroidal anti inflammatory drugs”), or two dispensings when at least one concerned the dispensing of a large pack size. Maternity concerned women between the ages of 15 to 49 years with 100% health insurance cover for maternity care from the first day of the 6th month of pregnancy until the twelfth day after delivery, or hospitalised during the year for delivery. “Hospitalisations for other reasons” concerned people with at least one hospital stay for a reason other than those taken into account for the diseases otherwise identified in the DHEM. In particular, this reason may be related to infection (pneumonia), trauma, surgery (hip prosthesis, appendicectomy), exploratory examinations (colonoscopy), or ill-defined symptoms or conditions.

Several health conditions may be identified in the same patient, but some conditions, corresponding to different states of the same disease, were exclusive during the same year (for example: acute coronary syndrome and chronic coronary heart disease; breast cancer during the active treatment phase and breast cancer under surveillance; chronic dialysis, renal transplantation and follow-up for renal transplantation). Similarly, chronic treatments and the corresponding diseases were exclusive, for example, “chronic psychoactive drug treatment (without a diagnosis of mental illness)”. The objective of algorithms identifying chronic treatments was to detect people who probably presented a certain disease, but without an ICD-10 code allowing identification of this disease.

### Expenditures

Reimbursed expenditures directly attributable to healthcare administered during a given year were calculated for each individual, from a national health insurance perspective. Expenditures funded by national health insurance, but not directly attributable to individuals (e.g. lump sums paid to healthcare institutions or healthcare professionals) and medical and social welfare expenditures were beyond the scope of this study. Three categories of expenditure were defined: ambulatory care, hospital care and cash benefits administered by national health insurance.

Ambulatory care expenditures comprised reimbursed expenditure related to healthcare administered by healthcare professionals, drugs, medical devices, laboratory tests, transport and other ambulatory care goods and services. These expenditures were derived directly from SNDS individual reimbursement data.

Hospital care expenditures comprised expenditures related to short-stay hospitalisations (“medicine, surgery, obstetrics” field of the hospital discharge database), rehabilitation care hospitalisations (“aftercare and rehabilitation” field), psychiatric hospitalisations, hospital at home and public hospital outpatient care. These expenditures were calculated for each stay from the specific expenditures of each diagnosis-related group.

Finally, cash benefits, also available in the SNDS for each individual, comprised daily allowances paid by national health insurance for sick leave or maternity leave and compensation for work accidents, occupational disease or loss of salary as a result of disability.

### Attribution of expenditures to health conditions

Reimbursed expenditures relative to short-stay, rehabilitation care or psychiatric hospitalisations and maternity cash benefits were directly attributed to the corresponding health conditions. A same stay could be attributed to only one health condition. As recommended in the methodology of the Health Expenditures by Diseases and Conditions project, priority was given to the principal diagnosis of the stay (with the ICD-10 codes of the health condition identification algorithms). (7) However, some very expensive diseases were considered to be responsible for the expenditure related to the stay, even when they were coded as a comorbid condition (haemophilia, hereditary metabolic diseases, amyloidosis, cystic fibrosis, dermatopolymyositis). Similarly, neonatal hospital stays were attributed directly to maternity care expenditure. Specific management rules were applied to the rare cases in which a stay was attributed to several health conditions. For stays that could not be attributed to a health condition, less disease-specific ICD-10 codes were used, based on the fact that the person had presented this health condition during the same year. Finally, short-stay hospitalisation expenditures not attributed to a health condition at the end of this process were attributed to “hospitalisations for other reasons”.

As the available information for all of the other types of expenditure was insufficient to directly attribute expenditures to health conditions, a top-down method was applied. For individuals with only one health condition identified in the year, expenditures were attributed to this health condition. For the other individuals, expenditures were distributed between health conditions *pro rata* to the mean expenditures calculated in individuals with only one health condition.

In order to avoid overestimating expenditures attributed to a given disease, a sum corresponding to the expenditures for so-called “usual” care was also subtracted from each type of ambulatory care expenditure for each individual, with the exception of transport and midwifery expenditures. The sum subtracted corresponded to the second decile of the expenditures of the age- and sex-matched population with healthcare utilisation corresponding to the type of expenditure considered.

Overall, the expenditure attributed to a health condition was therefore the sum of expenditures attributed by these various methods, with no possibility of double counting.

### Statistical analyses

Sociodemographic characteristics of the study population were described for 2012 and 2017, including healthcare reimbursement covered by complementary universal health insurance, attributed to low-income earners (€8,723 per year for a person living alone in metropolitan France in 2017) and then an indicator of social precariousness.

Crude annual prevalences of each health condition were calculated globally and by age (0 to 14 years-old, 15 to 34, 35 to 54, 55 to 64, 65 to 74, 75 or more) and sex, and were standardized for the age and sex structure of the French population at 01/01/2018 (source: Institut National de la Statistique et des Etudes Economiques https://www.insee.fr/fr/statistiques). The same analysis was performed for all people with at least one health condition. Total (€) and mean (€ per year and per patient) expenditures attributed to the various health conditions were then described, globally and by type of expenditure. Mean annual growth rates of expenditures and numbers of patients, the two components of total expenditure, were calculated for the period 2012-2017. In order to analyse expenditures according to categories of health conditions, we distinguished acute cardiovascular disease (CVD) from chronic CVD and cancers during the active treatment phase from cancers under surveillance, as these conditions correspond to very different situations in terms of management and costs.

Numbers were rounded off to the nearest thousand in view of the dimensions of the population and mean expenditures were rounded off to the nearest hundred euros. Proportions were expressed with two significant figures. As the analyses were not performed on a random sample of the population, no confidence interval was estimated. Statistical analyses were performed with SAS Enterprise Guide 4.3 software, SAS Institute Inc. Cary, NC.

## Results Population

In 2017, the DHEM comprised 57.6 million people (55.9 million in 2012), comprising 54% of women (54% in 2012). The median age was 40 years (interquartile range: 20-59), i.e., one year older than in 2012 (39 years (19-58)). The proportion of complementary universal health insurance beneficiaries was 9.6% (vs. 8.5% in 2012). The mortality rate was 7.7 per 1,000 people in 2017, versus 7.3 per 1,000 people in 2012.

### Prevalences of health conditions

In 2017, 45% (26.0 million) of the study population presented at least one health condition (Table 1). The most prevalent health conditions were “hospitalisations for other reasons” (14%), chronic antihypertensive treatment (without a diagnosis of cardiovascular disease, diabetes or end-stage renal disease (ESRD)) (11%), maternity (9.1%), diabetes (5.6%), chronic respiratory disease (5.5%), and chronic treatment with anxiolytics (4.8%) and antidepressants or mood regulators (4.5%) (without a diagnosis of mental illness). Prevalences standardized to the sex and age structure of the French population were very similar to crude prevalences.

**Table 1.** Crude and age- and sex-standardised prevalences of health conditions in 2017

|  | n, thousand | crude Pr. | standardised Pr.* |
| --- | --- | --- | --- |
| Whole population (thousand) | N=57,600 |  |  |
| At least one health condition | 25,967 | 45% | 45% |
| Cardiovascular diseases | 4016 | 7.00% | 7.30% |
| Acute cardiovascular diseases | 356 | 0.62% | 0.64% |
| Acute coronary syndrome | 81 | 0.14% | 0.15% |
| Acute stroke | 100 | 0.17% | 0.18% |
| Acute heart failure | 152 | 0.26% | 0.27% |
| Acute pulmonary embolism | 36 | 0.06% | 0.06% |
| Chronic cardiovascular diseases | 3897 | 6.80% | 7.10% |
| Chronic coronary heart disease | 1559 | 2.70% | 2.80% |
| Sequelae of stroke | 649 | 1.10% | 1.20% |
| Chronic heart failure | 520 | 0.90% | 0.94% |
| Peripheral vascular diseases | 597 | 1.00% | 1.10% |
| Arrhythmia or conduction disorders | 1357 | 2.40% | 2.40% |
| Valvular heart disease | 357 | 0.62% | 0.64% |
| Other cardiovascular diseases | 277 | 0.48% | 0.50% |
| Cardiovascular risk treatment<sup>a</sup> | 7307 | 12.70% | 13.00% |
| Antihypertensive treatment<sup>a</sup> | 6234 | 10.80% | 11.10% |
| Lipid-lowering treatment<sup>a</sup> | 2787 | 4.80% | 5.00% |
| Diabetes | 3237 | 5.60% | 5.80% |
| Cancers | 2637 | 4.60% | 4.70% |
| Cancers during the active treatment phase | 1189 | 2.10% | 2.10% |
| Female breast cancer | 193 | 0.62% | 0.64% |
| Colorectal cancer | 130 | 0.23% | 0.23% |
| Lung cancer | 80 | 0.14% | 0.14% |
| Prostate cancer | 170 | 0.64% | 0.65% |
| Other cancers | 671 | 1.20% | 1.20% |
| Follow-up for cancer | 1529 | 2.70% | 2.70% |
| Female breast cancer | 421 | 1.40% | 1.40% |
| Colorectal cancer | 165 | 0.29% | 0.30% |
| Lung cancer | 42 | 0.07% | 0.08% |
| Prostate cancer | 233 | 0.87% | 0.90% |
| Other cancers | 734 | 1.30% | 1.30% |
| Mental illness | 2190 | 3.80% | 3.90% |
| Psychotic disorders | 430 | 0.75% | 0.76% |
| Neurotic and mood disorders | 1199 | 2.10% | 2.10% |
| Mental impairment | 127 | 0.22% | 0.22% |
| Addictive disorders | 387 | 0.67% | 0.70% |
| Mental illness starting in childhood | 142 | 0.25% | 0.25% |
| Other mental illness | 410 | 0.71% | 0.72% |
| Chronic psychoactive drug treatment<sup>b</sup> | 5013 | 8.70% | 8.80% |
| Antidepressant or mood-regulating treatments<sup>b</sup> | 2598 | 4.50% | 4.50% |
| Neuroleptic treatments<sup>b</sup> | 286 | 0.50% | 0.51% |
| Anxiolytic treatments<sup>b</sup> | 2787 | 4.80% | 4.90% |
| Hypnotic treatments<sup>b</sup> | 1259 | 2.20% | 2.20% |
| Neurological or degenerative diseases | 1370 | 2.40% | 2.40% |
| Dementia | 613 | 1.10% | 1.10% |
| Parkinson disease | 215 | 0.37% | 0.39% |
| Multiple sclerosis | 98 | 0.17% | 0.17% |
| Paraplegia | 85 | 0.15% | 0.15% |
| Myopathy or myasthenia | 41 | 0.07% | 0.07% |
| Epilepsy | 292 | 0.51% | 0.51% |
| Other neurological conditions | 145 | 0.25% | 0.25% |
| Chronic respiratory diseases<sup>c</sup> | 3141 | 5.50% | 5.50% |
| Inflammatory or rare diseases or HIV infection | 1053 | 1.80% | 0.40% |
| Inflammatory bowel diseases | 227 | 0.39% | 0.40% |
| Rheumatoid arthritis and related diseases | 244 | 0.42% | 0.43% |
| Ankylosing spondylitis and related diseases | 182 | 0.32% | 0.32% |
| Other chronic inflammatory diseases | 173 | 0.30% | 0.30% |
| Hereditary metabolic diseases or amyloidosis | 99 | 0.17% | 0.18% |
| Cystic fibrosis | 8 | 0.01% | 0.01% |
| Haemophilia or severe coagulation disorders | 42 | 0.07% | 0.07% |
| HIV infection | 132 | 0.23% | 0.18% |
| End-stage renal disease | 82 | 0.14% | 0.15% |
| Chronic dialysis | 45 | 0.08% | 0.08% |
| Kidney transplant | 3 | 0.01% | 0.01% |
| Follow-up for kidney transplant | 34 | 0.06% | 0.06% |
| Liver or pancreatic diseases<sup>c</sup> | 512 | 0.89% | 0.92% |
| Other long-term diseases | 1479 | 2.60% | 0.39% |
| Maternity | 1222 | 9.10% | 8.90% |
| Hospitalisation for other reasons | 8052 | 14.00% | 0.24% |
| Analgesic or NSAID treatment<sup>d</sup> | 1346 | 2.30% | 2.30% |

\*standardisation according to sex and age based on estimations of the French population by *Institut National de la Statistique et des Etudes Economiques* on 01/01/2018.
\*\*prevalences among men or women only, as appropriate.
<sup>a</sup> without a diagnosis of cardiovascular disease, diabetes or ESRD. <sup>b</sup> without a diagnosis of mental illness. <sup>c</sup> excluding cystic fibrosis. <sup>d</sup> excluding diseases, treatments, maternity care or hospitalisation. Abbreviations: ESRD, end-stage renal disease; NSAID, non steroidal anti inflammatory drugs

As expected, the proportion of people with at least one health condition increased markedly with age, 6.6% before the age of 15 to 90% after the age of 74 in women and 9.9% to 91%, respectively, in men (Supplemental Table 1). The proportion of people between the ages of 15 and 34 years with at least one health condition was twofold higher in women (22%) than in men (11%).

A cardiovascular disease was identified in 7.0% of individuals. In people over the age of 74, cardiovascular disease was identified in 31% of women and 46% of men, most commonly chronic coronary heart disease (8.9% and 20%, respectively) and arrhythmias and conduction disorders (14% in women and 19% in men). Cardiovascular risk treatment (without a diagnosis of cardiovascular disease, diabetes or ESRD) was identified more frequently in women (14%, 44% after the age of 74) than in men (11%, 32% after the age of 74), most commonly consisting of antihypertensive treatment. Diabetes was identified in 5.6% of individuals, more frequently in men (6.4%) than in women (4.9%). The proportion of people with a cancer (all sites combined) was 4.6%, comparable between men and women, but higher in women than in men between the age of 15 and 64. A mental illness was identified in 3.8% of individuals and chronic psychoactive drug treatment (without a diagnosis of mental illness) was identified in 8.7% of individuals (particularly after the age of 74: 32% of women and 21% of men). Psychotic disorders were less common in women (0.63%) than in men (0.89%), in contrast with neurotic and mood disorders (2.6% and 1.5%, respectively). Finally, maternity care concerned 12% of women aged 15-34 years and 5.5% of women aged 35-54 years.

The prevalence of most health conditions increased with age. However, mental illness starting in childhood was more common among the youngest people, particularly boys. Multiple sclerosis was more common in women between the ages of 35 and 64. Cystic fibrosis was more common among young people. The prevalences of chronic respiratory diseases (combining asthma and chronic obstructive pulmonary disease) and “hereditary metabolic diseases and amyloidosis” were higher in individuals aged 0-14 years than in individuals aged 15-34 years, then increased again among older individuals. This pattern was also observed for “hospitalisations for other reasons”.

The age- and sex-standardized prevalences of “hospitalisations for other reasons” increased between 2012 and 2017 (especially in 2016), as did those of CVD, diabetes and mental illness (Figure 1). Inversely, these prevalences decreased for cardiovascular risk treatment (without a diagnosis of cardiovascular disease, diabetes or ESRD), chronic psychoactive drug treatment (without a diagnosis of mental illness) and maternity care.

**Figure 1.**
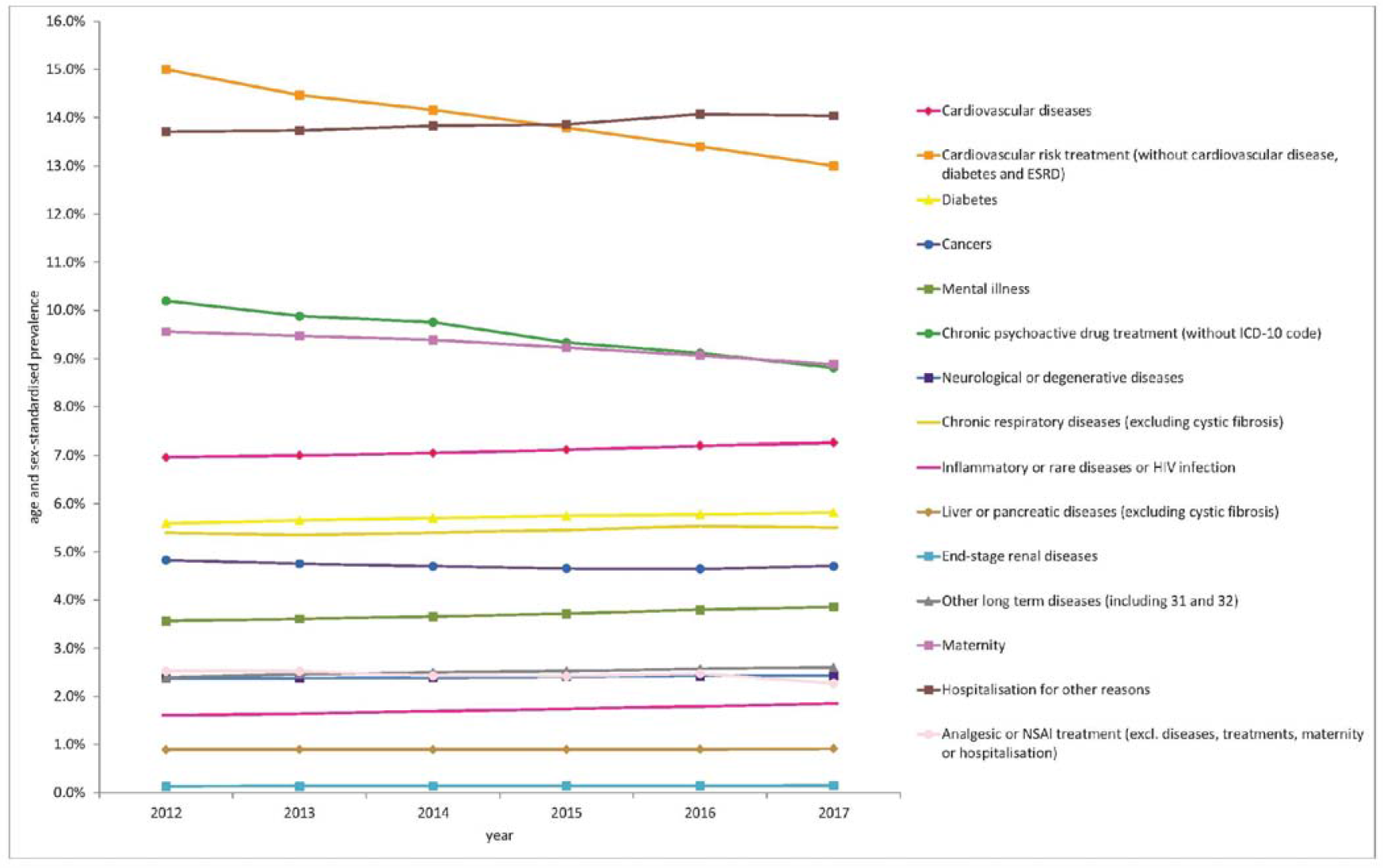
Variations in the prevalence of categories of health conditions between 2012 and 2017. Standardized prevalences according to the age and sex structure of the French population at 01/01/2018 (source: Institut National de la Statistique et des Etudes Economiques https://www.insee.fr/fr/statistiques).

### Reimbursed expenditures attributed to health conditions in 2017

In 2017, €140.1 billion were reimbursed by national health insurance for the healthcare of patients included in the DHEM (Table 2), versus €123.0 billion in 2012. “Hospitalisations for other reasons” accounted for the highest expenditure (€31.3 billion, i.e., 22% of total expenditure), followed by cancers, mental illness, CVD, maternity and diabetes, as detailed below.

**Table 2.** Distribution of total reimbursed expenditures (€140.1 billion) by health condition in 2017

|  | Reimbursed expenditures |  |
| --- | --- | --- |
|  | Total (in billion €) | Proportion |
| Cardiovascular diseases | 14.00 | 10% |
| Acute cardiovascular diseases | 3.45 | 2.5% |
| Acute coronary syndrome | 0.82 | 0.58% |
| Acute stroke | 1.26 | 0.90% |
| Acute heart failure | 1.20 | 0.85% |
| Acute pulmonary embolism | 0.18 | 0.13% |
| Chronic cardiovascular diseases | 10.55 | 7.5% |
| Chronic coronary heart disease | 2.76 | 2.0% |
| Sequelae of stroke | 1.78 | 1.3% |
| Chronic heart failure | 1.12 | 0.80% |
| Peripheral vascular diseases | 1.55 | 1.1% |
| Arrhythmia or conduction disorders | 1.86 | 1.3% |
| Valvular heart disease | 1.00 | 0.71% |
| Other cardiovascular diseases | 0.48 | 0.35% |
| Cardiovascular risk treatment<sup>a</sup> | 4.84 | 3.5% |
| Antihypertensive treatment<sup>a</sup> | 3.56 | 2.5% |
| Lipid-lowering treatment<sup>a</sup> | 1.28 | 0.91% |
| Diabetes | 6.99 | 5.0% |
| Cancers | 15.59 | 11% |
| Cancers during the active treatment phase | 13.97 | 10% |
| Female breast cancer | 2.40 | 1.7% |
| Colorectal cancer | 1.43 | 1.0% |
| Lung cancer | 1.60 | 1.1% |
| Prostate cancer | 1.07 | 0.76% |
| Other cancers | 7.47 | 5.3% |
| Follow-up for cancer | 1.62 | 1.2% |
| Female breast cancer | 0.48 | 0.34% |
| Colorectal cancer | 0.17 | 0.12% |
| Lung cancer | 0.07 | 0.05% |
| Prostate cancer | 0.16 | 0.12% |
| Other cancers | 0.74 | 0.53% |
| Mental illness | 14.53 | 10% |
| Psychotic disorders | 4.44 | 3.2% |
| Neurotic and mood disorders | 5.31 | 3.8% |
| Mental impairment | 0.57 | 0.41% |
| Addictive disorders | 1.39 | 0.99% |
| Mental illness starting in childhood | 1.19 | 0.85% |
| Other mental illness | 1.63 | 1.2% |
| Chronic psychoactive drug treatment<sup>b</sup> | 5.81 | 4.1% |
| Antidepressant or mood-regulating treatments<sup>b</sup> | 2.44 | 1.7% |
| Neuroleptic treatments<sup>b</sup> | 0.25 | 0.18% |
| Anxiolytic treatments<sup>b</sup> | 2.20 | 1.6% |
| Hypnotic treatments<sup>b</sup> | 0.93 | 0.66% |
| Neurological or degenerative diseases | 6.39 | 4.6% |
| Dementia | 2.08 | 1.5% |
| Parkinson disease | 0.85 | 0.61% |
| Multiple sclerosis | 1.13 | 0.81% |
| Paraplegia | 0.99 | 0.71% |
| Myopathy or myasthenia | 0.22 | 0.16% |
| Epilepsy | 0.51 | 0.36% |
| Other neurological conditions | 0.62 | 0.44% |
| Chronic respiratory diseases<sup>c</sup> | 2.95 | 2.1% |
| Inflammatory or rare diseases or HIV infection | 5.34 | 3.8% |
| Inflammatory bowel diseases | 0.85 | 0.61% |
| Rheumatoid arthritis and related diseases | 0.82 | 0.59% |
| Ankylosing spondylitis and related diseases | 0.79 | 0.56% |
| Other chronic inflammatory diseases | 0.37 | 0.26% |
| Hereditary metabolic diseases or amyloidosis | 0.47 | 0.34% |
| Cystic fibrosis | 0.27 | 0.20% |
| Haemophilia or severe coagulation disorders | 0.48 | 0.34% |
| HIV infection | 1.28 | 0.91% |
| End-stage renal disease | 3.49 | 2.5% |
| Chronic dialysis | 2.79 | 2.0% |
| Kidney transplant | 0.22 | 0.16% |
| Follow-up for kidney transplant | 0.47 | 0.34% |
| Liver or pancreatic diseases<sup>c</sup> | 1.79 | 1.3% |
| Other long-term diseases | 3.40 | 2.4% |
| Maternity | 7.75 | 5.5% |
| Hospitalisation for other reasons | 31.32 | 22% |
| Analgesic or NSAID treatment<sup>d</sup> | 14.00 | 10% |
| Usual Care | 5.19 | 3.7% |
| No health condition identified | 9.27 | 6.6% |

<sup>a</sup> without a diagnosis of cardiovascular disease, diabetes or ESRD. <sup>b</sup> without a diagnosis of mental illness. <sup>c</sup> excluding cystic fibrosis. <sup>d</sup> excluding diseases, treatments, maternity care or hospitalisation. Abbreviations: ESRD, end-stage renal disease; NSAID, non steroidal anti inflammatory drugs.

Expenditures attributed to the management of cancers (€15.6 billion, 11%) largely concerned cancers during the active treatment phase (€14.0 billion, 10% of total expenditure) (Table 2). Apart from the heterogeneous group of “other cancers during the active treatment phase”, breast cancer during the active treatment phase was the specific cancer site associated with the highest expenditure (€2.4 billion, 1.7% of total expenditure), ahead of lung cancer (€1.6 billion, 1.1%), colorectal cancer (€1.4 billion, 1.0%) and prostate cancer (€1.1 billion, 0.76%) during the active treatment phase. The predominant types of expenditure for cancers during the active treatment phase were short-stay hospitalisations (€7.1 billion) and pharmacy-dispensed drugs (€2.6 billion) (Figure 2).

**Figure 2.**
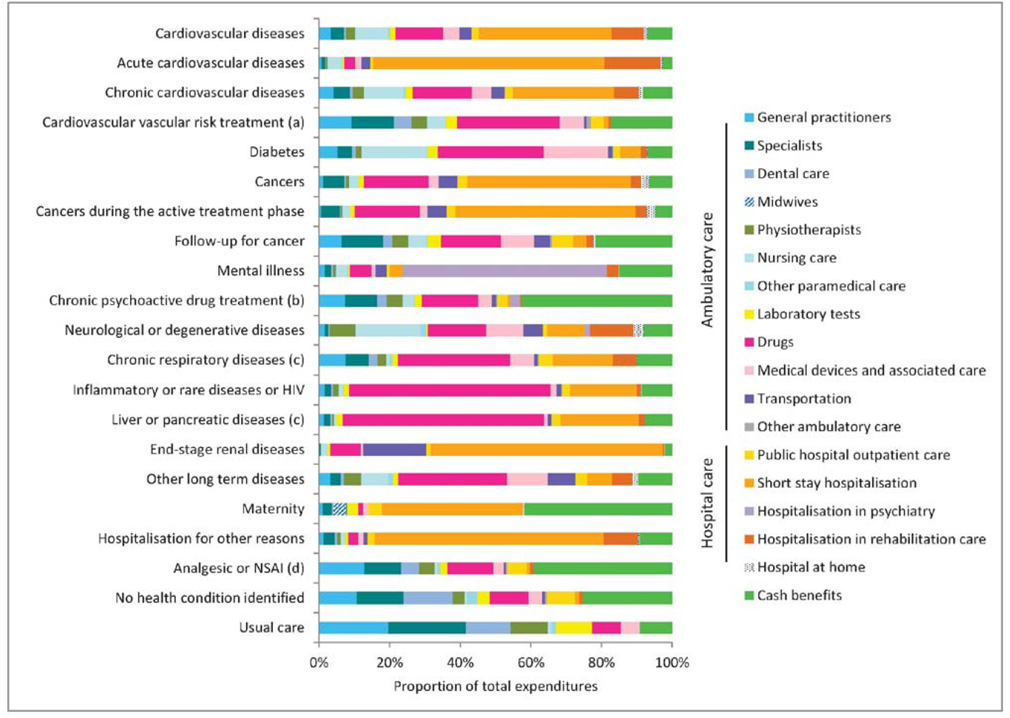
Distribution of reimbursed expenditures by type of expenditure according to health condition in 2017

Expenditures attributed to the management of mental illness (€14.5 billion, 10%) were primarily related to neurotic disorders and mood disorders (€5.3 billion, 3.8% of total expenditure) and psychotic disorders (€4.4 billion, 3.2%) (Table 2). Expenditures were largely related to psychiatric hospitalisations (€8.4 billion) (Figure 2).

Expenditures attributed to CVD (€14.0 billion, 10%) were essentially related to chronic CVD (€11.6 billion, 7.5% of total expenditure) (Table 2). Health conditions accounting for the largest share of expenditures were chronic coronary heart disease (€2.8 billion, 2.0% of total expenditure), arrhythmias or conduction disorders (€1.9 billion, 1.3%) and sequelae of stroke (€1.8 billion, 1.3%). Expenditures attributed to chronic CVD were essentially related to short-stay hospitalisations (€3.0 billion), pharmacy-dispensed drugs (€1.8 billion) and nursing care (€1.2 billion) (Figure 2). Stroke (€1.3 billion, 0.90% of total expenditure) and acute heart failure (€1.2 billion, 0.85%) were the conditions associated with the highest expenditures among the expenditures attributed to acute CVD (€3.5 billion, 2.5%). Expenditures attributed to acute CVD were essentially related to hospitalisations, mainly in short-stay wards (€2.3 billion) and rehabilitation care (€0.6 billion).

Expenditures attributed to maternity care represented €7.8 billion (5.5% of total expenditure). The main types of expenditures were short-stay hospitalisations (€3.1 billion) and cash benefits (€3.2 billion). Total midwife ambulatory care expenditure was €0.3 billion.

Finally, expenditures attributed to diabetes represented €7.0 billion (5.0% of total expenditure), with a large proportion corresponding to pharmacy-dispensed drugs (€2.1 billion), nursing care (€1.3 billion) and “other health products” (€1.3 billion), including medical devices.

For certain health conditions, especially cancers during the active treatment phase and acute CVD, the main determinant of total expenditure was the high mean expenditure per patient (Figure 3). Inversely, the main determinant of total expenditure for “hospitalisations for other reasons” and chronic psychoactive drug treatment (without a diagnosis of mental illness) and cardiovascular risk treatment (without a diagnosis of cardiovascular disease, diabetes or ESRD) was the large number of individuals concerned. ESRD was identified in only 82,000 individuals, but with an mean expenditure of €42,500 per patient.

**Figure 3.**
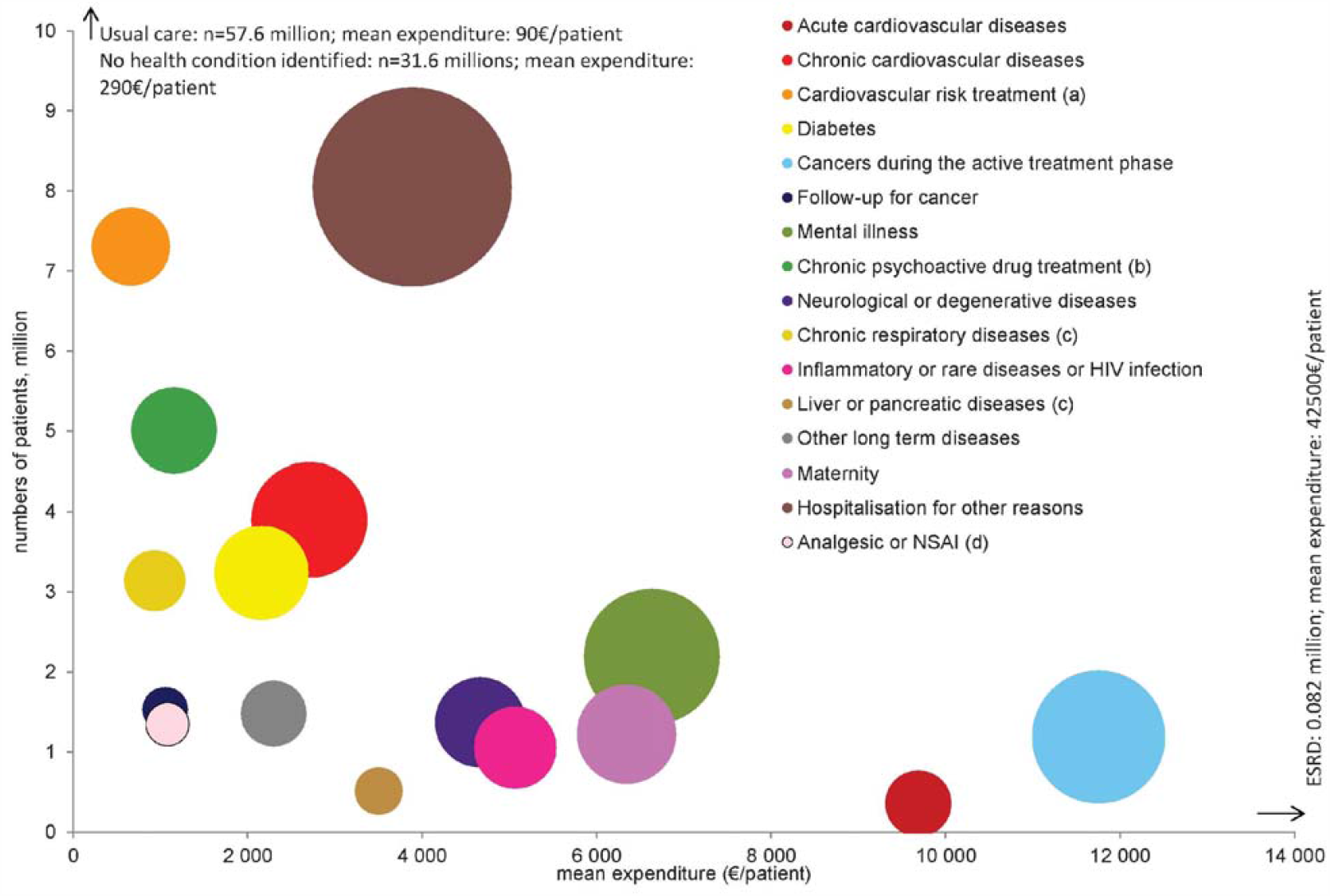
Sample sizes, mean reimbursed expenditures per year and per patient and total expenditures by category of health conditions in 2017. (a) without a diagnosis of cardiovascular disease, diabetes or end-stage renal disease (b) without ICD-10 code (from hospitalisations or long-term disease registration) (c) excluding cystic fibrosis (d) excluding diseases, treatments, maternity care or hospitalisation The size of bubbles is proportional to the total reimbursed expenditure. Bubbles for end-stage renal disease, people with no identified health condition and routine care were not represented because of very high mean expenditure and a very large sample size, respectively.

### Growth of reimbursed expenditures between 2012 and 2017

Between 2012 and 2017, healthcare expenditures increased by €17.1 billion (+14%). Expenditures increased for most health conditions, particularly “hospitalisations for other reasons” (+€3.2 billion, +2.3%/year), cancers during the active treatment phase (+€2.9 billion, +5.3%/year), mental illness (+€2.0 billion, +3.2%/year) and chronic CVD (+€1.6 billion, +3.6%/year) (Figure 4 and Figure 5). Expenditures for cardiovascular risk treatment (without a diagnosis of CVD, diabetes or ESRD) declined over this period (-€442 million, -1.7%/year).

**Figure 4.**
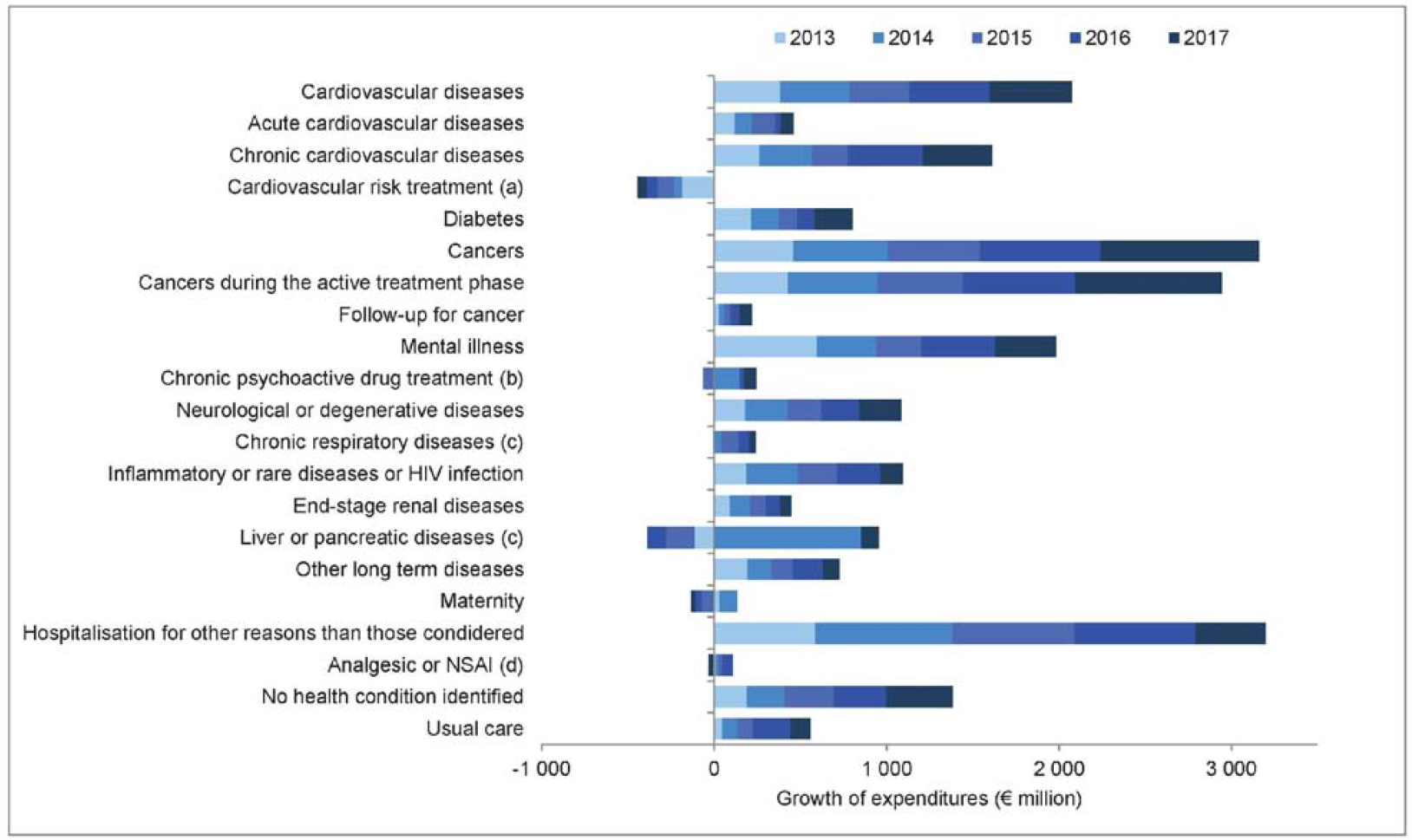
Annual growth of total reimbursed expenditures between 2012 and 2017 by category of health conditions. (a) without a diagnosis of cardiovascular disease, diabetes or end-stage renal disease (b) without ICD-10 code (from hospitalisations or long-term disease registration) (c) excluding cystic fibrosis (d) excluding diseases, treatments, maternity care or hospitalisation The values indicated are the difference of total reimbursed expenditures attributed to the health condition considered between years N and N-1.

**Figure 5.**
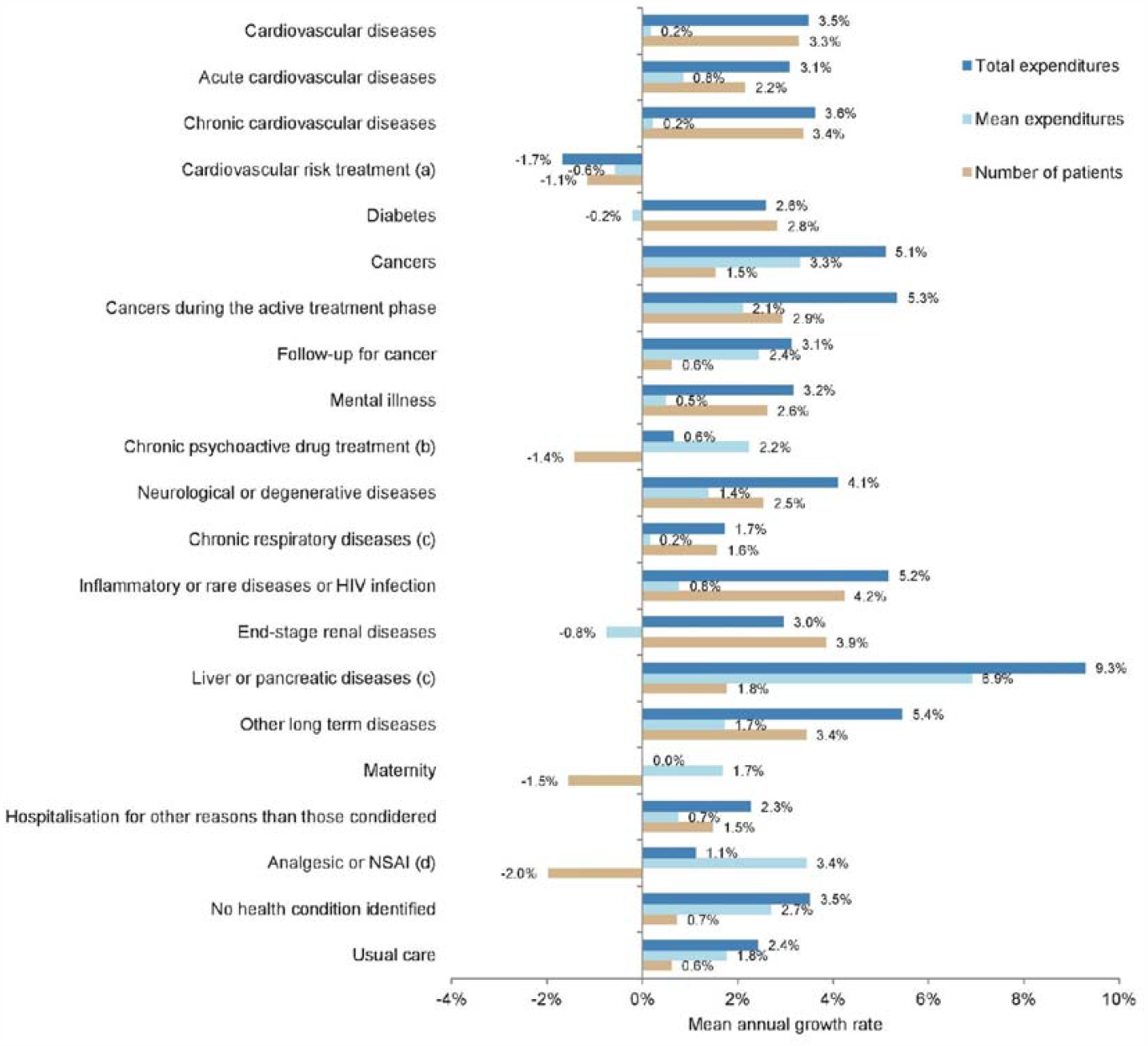
Mean annual growth rates, between 2012 and 2017, of total expenditures, number of patients and mean expenditures per patient by category of health conditions. (a) without a diagnosis of cardiovascular disease, diabetes or end-stage renal disease (b) without ICD-10 code (from hospitalisations or long-term disease registration) (c) excluding cystic fibrosis (d) excluding diseases, treatments, maternity care or hospitalisation

Despite a less marked absolute increase of expenditures, a high mean annual growth rate of total expenditures for liver and pancreatic disease was observed between 2012 and 2017 (+9.3%/year) (Figure 5). The largest share of this increase can be attributed to the increased mean expenditure per patient (+6.1%/year/patient). Expenditures attributed to liver or pancreatic disease increased considerably between 2013 and 2014 (+€852 million) (Figure 4). This growth was essentially related to drug expenditures (Supplemental Figure 1).

Similarly, for cancers during the active treatment phase, the largest share of the increased total expenditures (+5.3%/year) can be attributed to the increased mean expenditures (+3.1%/year/patient) (Figure 5). In particular, mean expenditures increased for prostate cancer (+9.0%/year/patient) (Supplemental Figure 2), related to drugs, specialist consultations and short-stay hospitalisations (Supplemental Figure 3). Mean expenditures attributed to colorectal cancer declined between 2012 and 2017 (−1.9%/year/patient), while the number of patients increased (+4.3%/year).

Inversely, the growth of total expenditure was essentially due to an increase of the number of patients for CVD (+3.3%/year), diabetes (+2.8%/year), mental illness (+2.6%/year), chronic respiratory diseases (+1.6%/year), ESRD (+3.9%/year) and the “rare or inflammatory diseases or HIV infection” category (+4.2%/year) (Figure 5). Within this last category of health conditions, the numbers of patients increased for the most prevalent diseases: +7.1%/year for ankylosing spondylitis and related diseases, +4.1%/year for rheumatoid arthritis and related diseases and +3.4%/year for chronic inflammatory bowel disease (Supplemental Figure 4). Furthermore, total expenditures attributed to cystic fibrosis, although a rare disease (prevalence: 0.014%, see Table 1), markedly increased (+19%/year), almost exclusively due to an increase of the mean expenditures per patient (+16%/year). This relative increase of total expenditures was the highest of all health conditions studied in the DHEM and was related to increased drug expenditures in 2016 (Supplemental Figure 5).

## Discussion

For the first time in France, the DHEM has allowed a nationwide dual epidemiological and economic approach, by identifying a large number of health conditions on the basis of medical and claims data and then by distributing all reimbursed expenditures between these health conditions, without double counting. The most prevalent diseases were diabetes, chronic respiratory diseases and chronic coronary heart disease. As expected, the highest expenditures were observed for cancers during the active treatment phase and chronic cardiovascular disease, but also mental illness. The most marked increase in total expenditures over the study period concerned liver or pancreatic diseases. Apart from diseases, maternity and, by construction, “hospitalisations for other reasons”, were among the most prevalent and most expensive health conditions.

A limitation of this study is that the information available in the SNDS could not be used to distinguish between certain health conditions, such as chronic obstructive pulmonary disease and asthma, and cannot be used to study people with no reimbursed healthcare utilization during the year. The prevalences of health conditions may have been underestimated in people with rare healthcare utilization, such as young men. (22) Inversely, the availability of very extensive individual data in the SNDS allowed detailed analysis and allowed the majority of expenditures to be directly attributed to health conditions. *Pro rata* estimation of ambulatory care expenditures for people with several health conditions was sometimes based on small number of patients, but the minor variations in these expenditures over time suggest that this method was robust. Finally, due to the nationwide coverage of the SNDS and the fact that national health insurance is mandatory in France, the DHEM allowed analysis of almost all of the population, avoiding a non-participation bias. Nevertheless, the results would probably be different for health insurance schemes not included in the DHEM (13% of the French population, mainly farmers and self-employed workers). In years to come, the DHEM will include all of the French population, but will no longer concern the years prior to 2015 due to missing LTD data for certain schemes. (16)

The main limitations of most of the studies conducted with a similar objective in other countries were related to missing data, concerning either individual data related to healthcare expenditures or ambulatory care medical diagnoses, skilled nursing home data, or the fact that the study was based on a limited population. (4,7–12,15,23) In France, diagnoses are not coded for each ambulatory care visit, but LTD diagnoses allowed identification of a large number of serious or expensive chronic diseases managed in the ambulatory care setting. Healthcare expenditures not related to individual patients are not included in the DHEM and were therefore not studied, including in skilled nursing homes, especially for the third of these nursing homes funded by lump sum payments. The DHEM included a greater number of health conditions than in many previous studies (7–9,11,12). However, no specific algorithm is available to identify certain musculoskeletal disorders, such as osteoarthritis and low back and neck pain other than by “chronic treatment with analgesics or non steroidal anti inflammatory drugs”, although several studies have reported these conditions to be responsible for high expenditures. (8–10)

Comparisons of international results are complex due to differences in health systems and health insurance cover, technical, methodological, epidemiological and demographic differences. Nevertheless, the main results concerning the most prevalent and most expensive diseases are consistent with those of similar studies conducted in other countries. Differences concerned the higher proportion of expenditures related to cancers in France (11%) than in most other countries (between 5.5% in the USA and 10% in the Czech Republic), except in Hungary (13%), Greece (13%) and Finland (12%) (7,10) and the share of expenditures for maternity care (5.5%), higher than in the USA (2.6%) (10) and Switzerland (1.1%) (8), probably because of maternity leave and a higher birth rate in France than in Switzerland (source: CIA https://www.cia.gov/library/publications/the-world-factbook/rankorder/2054rank.html#by).

The results of this study suggest several explanations for the growth of expenditures attributed to health conditions. The regular growth of expenditures for CVD and diabetes, essentially related to the growing number of patients, can probably be explained by ageing of the population. For other health conditions, the growth of total expenditures was mainly related to the growth of mean expenditures per patient. Therapeutic innovations coincided, in certain years, with a very marked growth of drug expenditures for liver and pancreatic diseases (new direct-acting antivirals against the hepatitis C virus in 2014) and cystic fibrosis (lumacaftor-ivacaftor combination, new indication for ivacaftor in 2015). The high but more regular growth of mean expenditures attributed to prostate cancer can probably be partly explained by the increased use of endocrine therapy and oral targeted therapies. The decline in mean expenditure attributed to colorectal cancer and the increased number of patients can be explained by the lower cost of several drugs during the study period and the use of a new screening test introduced in 2015.

The data used to identify health conditions and individual calculated expenditures are accessible via the SNDS to a large number of health system actors, which facilitates research on health services and care pathways, health economics studies and territorial diagnoses allowing local adaptation of public health policies. Each year, the DHEM is also used to enlighten health policies, particularly the public health act and the social security funding bill. One of the main results for political decision-makers was the prevalence and expenditures attributed to mental illness and psychoactive drug treatments, that had previously been underestimated. Similar findings have been reported in many other countries (7, 8,12, 23). Our results therefore provided a basis for action proposals to parliament, especially concerning the creation of specific healthcare provision to improve the management of somatic comorbidities, specifically devoted to people with serious mental illness, and to improve the relevance of psychoactive drug treatments. (24) An experimentation to evaluate the efficacy of psychotherapy as a replacement for drug treatment for non-serious depression has also been initiated.

Future studies could assess the performances of the algorithms used to identify health conditions on the basis of medical administrative data, that have been rarely studied in the world up until now, due to the lack of gold standards. Clinical data derived from the CONSTANCES general population cohort were used to evaluate diabetes identification algorithms in the SNDS, with very good performances for the algorithms studied (including that used in the DHEM). (25,26) This type of cohort could be used more systematically in the future to evaluate other disease identification algorithms. The use of predictive models to identify diseases is also an alternative to algorithms that has not been extensively studied to date. (27) Another line of research concerns the methods of attribution of expenditures to health conditions. In countries in which individual data are available, expenditure modelling could more reliably take into account the interactions between health conditions and expenditures. (9)

## Data Availability

Data cannot be shared publicly because it is forbidden by law (sensitive individual
data). Data are available from the Health Data Hub (contact via hdh@health-datahub.
fr) for researchers who meet the criteria for access to confidential data.

## Acknowledgements

The authors are very grateful to Anne Cuerq, Marjorie Mazars, Alexandre Rigault, Solène Sansom and Stéphane Tala for their contribution to the development of the DHEM, Anne-Sophie Aguadé for her support and Moussa Laanani for reviewing this paper prior to submission.

## Supplemental Material

**Supplemental Table 1.** Prevalences of health conditions by sex and age in 2017

|  | women |  |  |  |  |  |  | men |  |  |  |  |  |  |
| --- | --- | --- | --- | --- | --- | --- | --- | --- | --- | --- | --- | --- | --- | --- |
|  | all ages | 0-14 | 15-34 | 35-54 | 55-64 | 65-74 | 75+ | all ages | 0-14 | 15-34 | 35-54 | 55-64 | 65-74 | 75+ |
| Whole population (thousand) | 30,928 | 5,237 | 7,440 | 8,011 | 3,827 | 3,234 | 3,179 | 26,672 | 5,489 | 6,538 | 6,895 | 3,191 | 2,654 | 1,905 |
| At least one health condition | 48 | 20 | 31 | 44 | 64 | 78 | 93 | 42 | 24 | 18 | 37 | 66 | 82 | 93 |
| Cardiovascular diseases | 5.5 | 0.28 | 0.45 | 1.7 | 5.2 | 10 | 31 | 8.7 | 0.34 | 0.56 | 3.6 | 14 | 26 | 46 |
| Acute cardiovascular diseases | 0.55 | <0.01 | 0.031 | 0.14 | 0.36 | 0.79 | 3.6 | 0.70 | <0.01 | 0.036 | 0.34 | 0.98 | 1.7 | 4.4 |
| Acute coronary syndrome | 0.086 | <0.01 | <0.01 | 0.036 | 0.10 | 0.17 | 0.45 | 0.21 | <0.01 | <0.01 | 0.17 | 0.41 | 0.53 | 0.80 |
| Acute stroke | 0.16 | <0.01 | 0.012 | 0.057 | 0.12 | 0.26 | 0.99 | 0.19 | <0.01 | 0.014 | 0.090 | 0.27 | 0.49 | 1.1 |
| Acute heart failure | 0.26 | <0.01 | <0.01 | 0.020 | 0.087 | 0.27 | 2.0 | 0.27 | <0.01 | <0.01 | 0.047 | 0.23 | 0.59 | 2.4 |
| Acute pulmonary embolism | 0.063 | <0.01 | 0.013 | 0.031 | 0.055 | 0.12 | 0.31 | 0.062 | <0.01 | 0.010 | 0.043 | 0.10 | 0.16 | 0.28 |
| Chronic cardiovascular diseases | 5.3 | 0.28 | 0.43 | 1.6 | 5.0 | 10 | 30 | 8.5 | 0.33 | 0.53 | 3.4 | 14 | 25 | 45 |
| Chronic coronary heart disease | 1.6 | <0.01 | 0.019 | 0.40 | 1.7 | 3.4 | 8.9 | 4.0 | <0.01 | 0.045 | 1.5 | 7.1 | 13 | 20 |
| Sequelae of stroke | 1.0 | 0.047 | 0.10 | 0.47 | 1.1 | 1.8 | 5.3 | 1.2 | 0.065 | 0.12 | 0.57 | 1.9 | 3.4 | 6.8 |
| Chronic heart failure | 0.87 | 0.024 | 0.021 | 0.11 | 0.44 | 1.1 | 6.4 | 0.95 | 0.025 | 0.031 | 0.25 | 1.1 | 2.3 | 7.1 |
| Peripheral vascular diseases | 0.64 | <0.01 | 0.014 | 0.16 | 0.70 | 1.2 | 3.7 | 1.5 | <0.01 | 0.016 | 0.41 | 2.8 | 5.0 | 7.8 |
| Arrhythmia or conduction disorders | 2.1 | 0.058 | 0.17 | 0.40 | 1.2 | 3.5 | 14 | 2.7 | 0.069 | 0.20 | 0.70 | 2.8 | 7.2 | 19 |
| Valvular heart disease | 0.57 | 0.015 | 0.033 | 0.13 | 0.42 | 1.0 | 3.5 | 0.68 | 0.018 | 0.045 | 0.22 | 0.79 | 1.8 | 4.6 |
| Other cardiovascular diseases | 0.37 | 0.16 | 0.10 | 0.17 | 0.39 | 0.67 | 1.5 | 0.61 | 0.18 | 0.13 | 0.27 | 0.87 | 1.7 | 2.8 |
| Cardiovascular risk treatment<sup>a</sup> | 14 | 0.044 | 0.86 | 8.7 | 26 | 39 | 44 | 11 | 0.057 | 0.73 | 8.8 | 25 | 32 | 32 |
| Antihypertensive treatment<sup>a</sup> | 12 | 0.038 | 0.78 | 7.7 | 21 | 32 | 40 | 9.1 | 0.051 | 0.63 | 7.1 | 20 | 27 | 28 |
| Lipid-lowering treatment<sup>a</sup> | 5.1 | <0.01 | 0.091 | 1.6 | 9.2 | 18 | 17 | 4.5 | <0.01 | 0.12 | 3.0 | 10 | 15 | 14 |
| Diabetes | 4.9 | 0.13 | 0.56 | 2.6 | 8.4 | 13 | 16 | 6.4 | 0.13 | 0.54 | 3.6 | 13 | 21 | 23 |
| Cancers | 4.6 | 0.10 | 0.56 | 3.3 | 7.4 | 11 | 14 | 4.5 | 0.12 | 0.36 | 1.6 | 6.6 | 15 | 25 |
| Cancers during the active treatment phase | 1.9 | 0.050 | 0.27 | 1.5 | 3.0 | 4.6 | 5.5 | 2.3 | 0.057 | 0.17 | 0.813 | 3.6 | 7.6 | 12 |
| Female breast cancer | 0.62 | <0.01 | 0.036 | 0.67 | 1.2 | 1.6 | 1.3 |  |  |  |  |  |  |  |
| Colorectal cancer | 0.19 | <0.01 | <0.01 | 0.084 | 0.30 | 0.54 | 0.72 | 0.26 | <0.01 | <0.01 | 0.093 | 0.47 | 0.96 | 1.2 |
| Lung cancer | 0.094 | <0.01 | <0.01 | 0.051 | 0.22 | 0.28 | 0.22 | 0.19 | <0.01 | <0.01 | 0.071 | 0.43 | 0.72 | 0.67 |
| Prostate cancer |  |  |  |  |  |  |  | 0.64 | <0.01 | <0.01 | 0.064 | 0.82 | 2.4 | 4.0 |
| Other cancers | 1.0 | 0.049 | 0.22 | 0.70 | 1.5 | 2.4 | 3.6 | 1.3 | 0.057 | 0.16 | 0.60 | 2.1 | 3.9 | 6.6 |
| Follow-up for cancer | 2.8 | 0.050 | 0.29 | 1.8 | 4.6 | 7.2 | 9.4 | 2.4 | 0.059 | 0.19 | 0.78 | 3.2 | 7.9 | 14 |
| Female breast cancer | 1.4 | <0.01 | 0.016 | 0.81 | 2.5 | 3.9 | 4.2 |  |  |  |  |  |  |  |
| Colorectal cancer | 0.27 | <0.01 | <0.01 | 0.075 | 0.32 | 0.66 | 1.3 | 0.31 | <0.01 | <0.01 | 0.073 | 0.40 | 1.0 | 2.0 |
| Lung cancer | 0.049 | <0.01 | <0.01 | 0.021 | 0.11 | 0.15 | 0.14 | 0.10 | <0.01 | <0.01 | 0.024 | 0.18 | 0.38 | 0.46 |
| Prostate cancer |  |  |  |  |  |  |  | 0.87 | <0.01 | <0.01 | 0.026 | 0.79 | 3.3 | 6.2 |
| Other cancers | 1.3 | 0.049 | 0.27 | 0.97 | 1.8 | 2.8 | 4.2 | 1.3 | 0.058 | 0.19 | 0.66 | 1.9 | 3.6 | 6.5 |
| Mental illness | 3.9 | 0.65 | 2.0 | 4.3 | 6.2 | 5.6 | 8.1 | 3.7 | 1.5 | 2.7 | 5.1 | 5.6 | 4.2 | 4.9 |
| Psychotic disorders | 0.63 | <0.01 | 0.32 | 0.87 | 1.1 | 0.94 | 0.92 | 0.89 | 0.020 | 0.82 | 1.6 | 1.2 | 0.79 | 0.52 |
| Neurotic and mood disorders | 2.6 | 0.10 | 1.1 | 2.9 | 4.3 | 3.9 | 6.1 | 1.5 | 0.10 | 0.84 | 2.1 | 2.7 | 2.0 | 2.7 |
| Mental impairment | 0.18 | 0.13 | 0.16 | 0.24 | 0.28 | 0.18 | 0.075 | 0.26 | 0.22 | 0.25 | 0.33 | 0.36 | 0.20 | 0.077 |
| Addictive disorders | 0.39 | <0.01 | 0.28 | 0.58 | 0.69 | 0.51 | 0.32 | 1.0 | <0.01 | 0.74 | 1.7 | 1.8 | 1.2 | 0.76 |
| Mental illness starting in childhood | 0.14 | 0.39 | 0.20 | 0.057 | 0.038 | 0.022 | 0.023 | 0.37 | 1.1 | 0.43 | 0.10 | 0.054 | 0.023 | 0.021 |
| Other mental illness | 0.77 | 0.077 | 0.56 | 0.82 | 1.0 | 1.0 | 1.7 | 0.65 | 0.13 | 0.52 | 0.85 | 0.87 | 0.76 | 1.4 |
| Chronic psychoactive drug treatment<sup>b</sup> | 11 | 0.10 | 2.8 | 11 | 17 | 21 | 32 | 6.1 | 0.21 | 1.8 | 6.4 | 10 | 13 | 20 |
| Antidepressant or mood-regulating treatments<sup>b</sup> | 6.0 | 0.024 | 1.8 | 7.1 | 9.8 | 10 | 14 | 2.8 | 0.032 | 0.99 | 3.6 | 5.0 | 4.9 | 8.0 |
| Neuroleptic treatments<sup>b</sup> | 0.49 | 0.028 | 0.15 | 0.41 | 0.60 | 0.68 | 2.0 | 0.50 | 0.12 | 0.31 | 0.58 | 0.66 | 0.65 | 1.4 |
| Anxiolytic treatments<sup>b</sup> | 6.1 | 0.057 | 1.5 | 5.7 | 9.2 | 12 | 18 | 3.4 | 0.075 | 0.95 | 3.6 | 5.9 | 7.1 | 11 |
| Hypnotic treatments<sup>b</sup> | 2.7 | <0.01 | 0.32 | 1.9 | 4.0 | 5.9 | 9.5 | 1.6 | <0.01 | 0.24 | 1.3 | 2.9 | 4.2 | 6.7 |
| Neurological or degenerative diseases | 2.6 | 0.44 | 0.74 | 1.3 | 1.8 | 2.8 | 15 | 2.1 | 0.53 | 0.84 | 1.4 | 2.1 | 3.4 | 12 |
| Dementia | 1.4 | <0.01 | <0.01 | 0.030 | 0.17 | 0.89 | 12 | 0.71 | <0.01 | <0.01 | 0.045 | 0.23 | 0.97 | 8.0 |
| Parkinson disease | 0.36 | <0.01 | <0.01 | 0.055 | 0.25 | 0.75 | 2.2 | 0.39 | <0.01 | <0.01 | 0.061 | 0.33 | 1.1 | 3.2 |
| Multiple sclerosis | 0.23 | <0.01 | 0.13 | 0.41 | 0.42 | 0.29 | 0.12 | 0.10 | <0.01 | 0.055 | 0.18 | 0.18 | 0.14 | 0.059 |
| Paraplegia | 0.12 | 0.030 | 0.068 | 0.14 | 0.20 | 0.20 | 0.18 | 0.18 | 0.038 | 0.11 | 0.22 | 0.28 | 0.29 | 0.27 |
| Myopathy or myasthenia | 0.069 | 0.020 | 0.044 | 0.082 | 0.094 | 0.10 | 0.11 | 0.076 | 0.036 | 0.061 | 0.079 | 0.10 | 0.11 | 0.13 |
| Epilepsy | 0.45 | 0.30 | 0.37 | 0.42 | 0.48 | 0.49 | 0.87 | 0.57 | 0.35 | 0.45 | 0.61 | 0.76 | 0.71 | 0.98 |
| Other neurological conditions | 0.23 | 0.12 | 0.17 | 0.23 | 0.32 | 0.33 | 0.40 | 0.27 | 0.14 | 0.21 | 0.26 | 0.39 | 0.42 | 0.50 |
| Chronic respiratory diseases<sup>c</sup> | 5.2 | 4.1 | 2.5 | 4.3 | 7.0 | 8.3 | 11 | 5.7 | 6.3 | 2.3 | 3.6 | 7.2 | 10 | 14 |
| Inflammatory or rare diseases or HIV infection | 2.0 | 0.26 | 0.98 | 2.4 | 3.0 | 3.2 | 3.7 | 1.6 | 0.28 | 0.88 | 2.1 | 2.8 | 2.8 | 3.1 |
| Inflammatory bowel diseases | 0.40 | 0.022 | 0.40 | 0.61 | 0.52 | 0.41 | 0.34 | 0.39 | 0.024 | 0.36 | 0.53 | 0.58 | 0.56 | 0.45 |
| Rheumatoid arthritis and related diseases | 0.58 | 0.045 | 0.11 | 0.46 | 1.0 | 1.3 | 1.6 | 0.24 | 0.020 | 0.037 | 0.16 | 0.44 | 0.64 | 0.91 |
| Ankylosing spondylitis and related diseases | 0.31 | 0.010 | 0.15 | 0.50 | 0.56 | 0.43 | 0.28 | 0.32 | <0.01 | 0.15 | 0.48 | 0.58 | 0.55 | 0.48 |
| Other chronic inflammatory diseases | 0.42 | 0.039 | 0.14 | 0.40 | 0.58 | 0.77 | 1.2 | 0.16 | 0.047 | 0.042 | 0.11 | 0.21 | 0.37 | 0.73 |
| Hereditary metabolic diseases or amyloidosis | 0.15 | 0.10 | 0.085 | 0.13 | 0.19 | 0.27 | 0.27 | 0.20 | 0.11 | 0.090 | 0.16 | 0.31 | 0.41 | 0.41 |
| Cystic fibrosis | 0.012 | 0.022 | 0.021 | <0.01 | <0.01 | <0.01 | <0.01 | 0.015 | 0.023 | 0.027 | 0.011 | <0.01 | <0.01 | <0.01 |
| Haemophilia or severe coagulation disorders | 0.071 | 0.014 | 0.041 | 0.086 | 0.10 | 0.11 | 0.13 | 0.075 | 0.041 | 0.050 | 0.073 | 0.10 | 0.13 | 0.15 |
| HIV infection | 0.15 | 0.011 | 0.083 | 0.34 | 0.21 | 0.084 | 0.029 | 0.32 | 0.010 | 0.16 | 0.65 | 0.66 | 0.29 | 0.11 |
| End-stage renal disease | 0.11 | <0.01 | 0.026 | 0.093 | 0.18 | 0.24 | 0.28 | 0.18 | <0.01 | 0.041 | 0.16 | 0.31 | 0.46 | 0.67 |
| Chronic dialysis | 0.059 | <0.01 | <0.01 | 0.031 | 0.076 | 0.14 | 0.24 | 0.10 | <0.01 | 0.010 | 0.050 | 0.14 | 0.27 | 0.58 |
| Kidney transplant | <0.01 | <0.01 | <0.01 | <0.01 | <0.01 | <0.01 | <0.01 | <0.01 | <0.01 | <0.01 | 0.011 | 0.014 | 0.016 | <0.01 |
| Follow-up for kidney transplant | 0.044 | <0.01 | 0.016 | 0.056 | 0.091 | 0.095 | 0.035 | 0.076 | <0.01 | 0.027 | 0.10 | 0.16 | 0.18 | 0.085 |
| Liver or pancreatic diseases<sup>c</sup> | 0.73 | 0.041 | 0.28 | 0.71 | 1.3 | 1.5 | 1.5 | 1.1 | 0.042 | 0.28 | 1.3 | 2.3 | 2.3 | 2.1 |
| Other long-term diseases | 2.8 | 1.1 | 1.4 | 1.8 | 3.2 | 4.5 | 9.6 | 2.2 | 1.3 | 1.4 | 1.5 | 2.8 | 4.1 | 7.0 |
| Maternity | 9.1 | 0 | 12 | 5.5 | 0 | 0 | 0 |  |  |  |  |  |  |  |
| Hospitalisation for other reasons | 14 | 9.9 | 9.2 | 13 | 16 | 21 | 27 | 14 | 12 | 8.4 | 11 | 17 | 23 | 30 |
| Analgesic or NSAID treatment<sup>d</sup> | 2.7 | 4.2 | 2.4 | 3.4 | 2.3 | 1.6 | 1.0 | 1.9 | 3.9 | 0.98 | 2.0 | 1.4 | 0.87 | 0.59 |

Data are % unless otherwise specified. <sup>a</sup> without a diagnosis of cardiovascular disease, diabetes or ESRD. <sup>b</sup> without a diagnosis of mental illness. <sup>c</sup> excluding cystic fibrosis. <sup>d</sup> excluding diseases, treatments, maternity care or hospitalisation. Abbreviations: ESRD, end-stage renal disease; NSAID, non steroidal anti inflammatory drugs

**Supplemental Figure 1.**
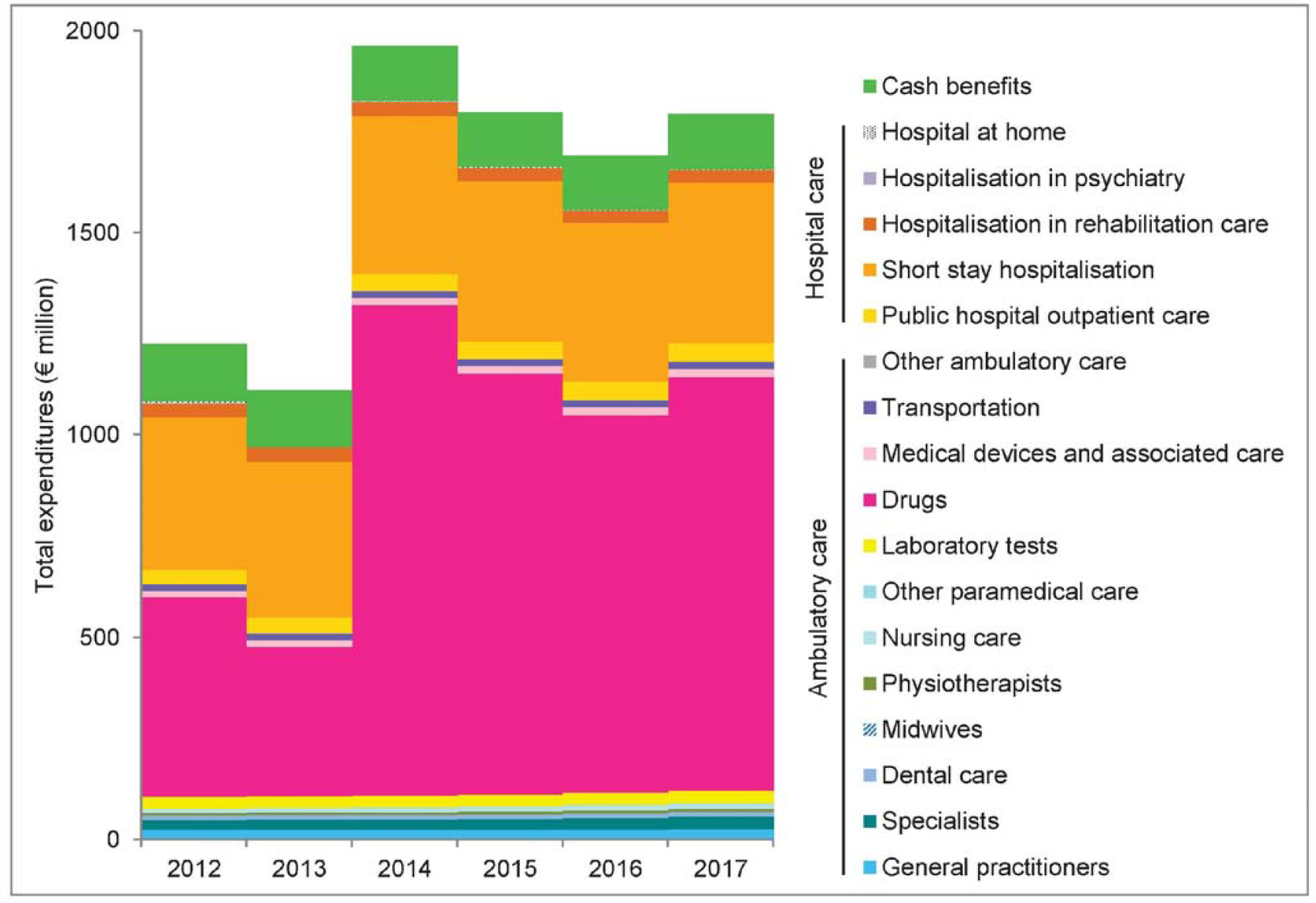
Growth of total expenditure attributed to liver and pancreatic diseases by type of expenditure between 2012 and 2017

**Supplemental Figure 2.**
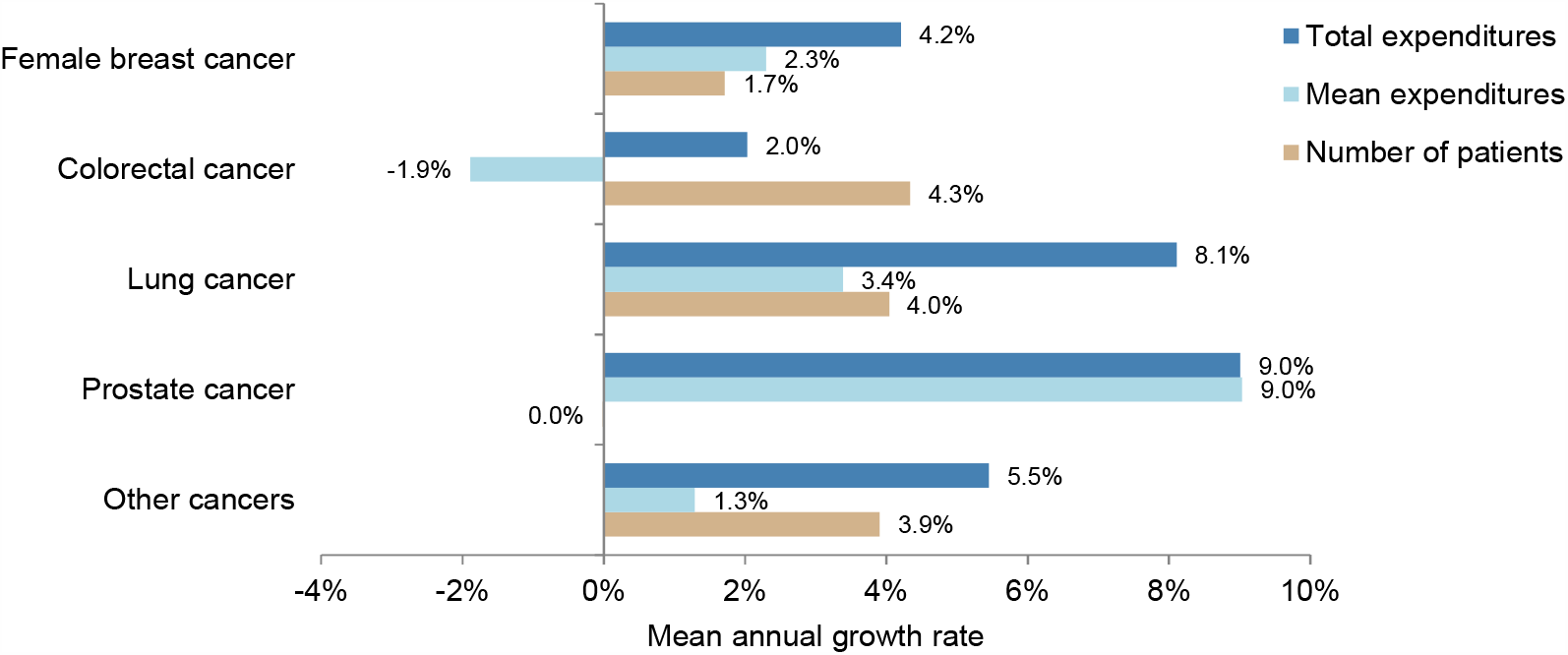
Mean annual growth rates between 2012 and 2017 of total expenditures, number of patients and mean expenditures per patient by health condition of the “cancers during the active treatment phase” category.

**Supplemental Figure 3.**
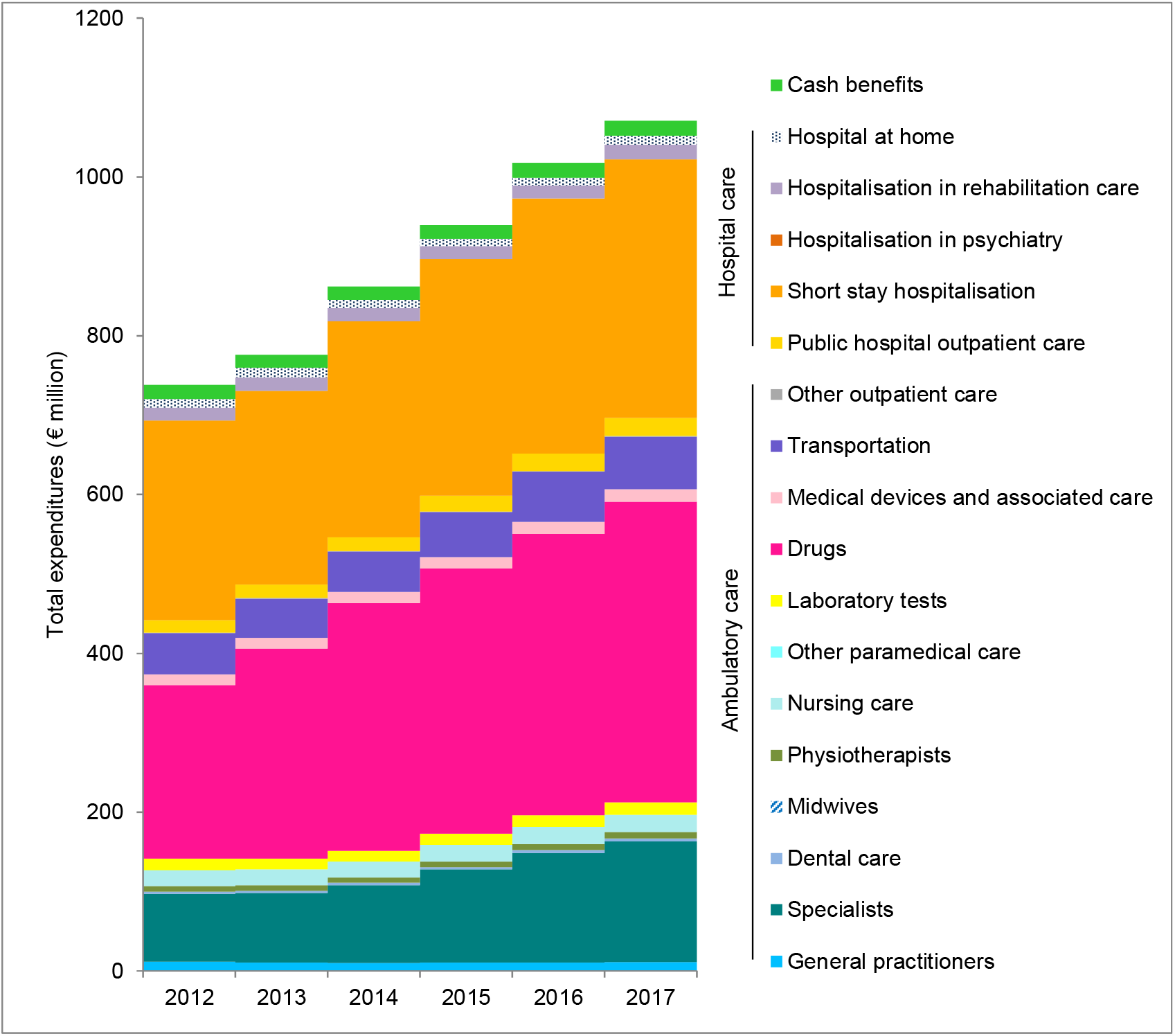
Growth of total expenditures attributed to prostate cancer during the active treatment phase by type of expenditure between 2012 and 2017.

**Supplemental Figure 4.**
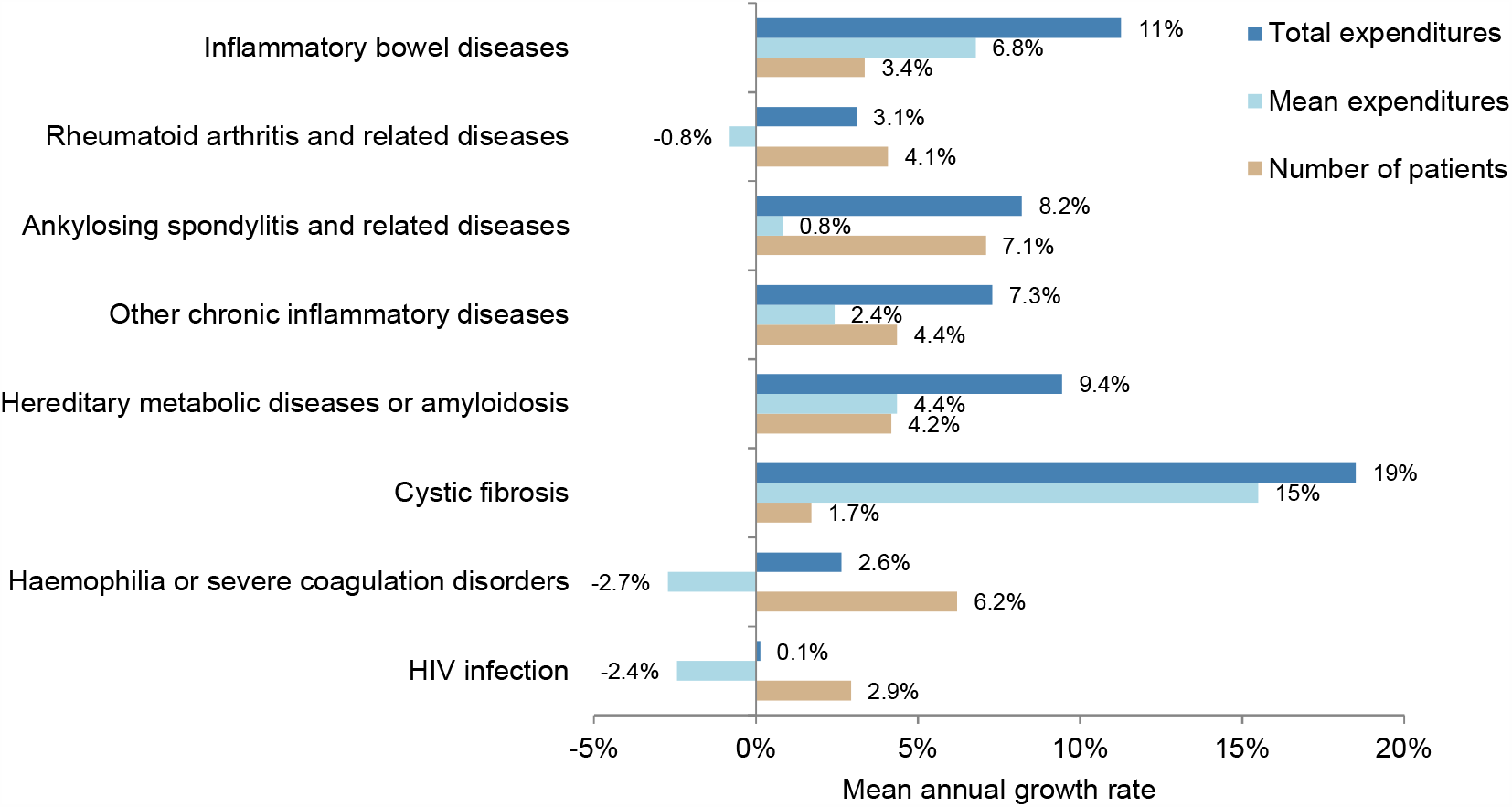
Mean annual growth rates between 2012 and 2017 of total expenditures, number of patients and mean expenditures per patient by health conditions of the “rare or inflammatory diseases or HIV infection” category.

**Supplemental Figure 5.**
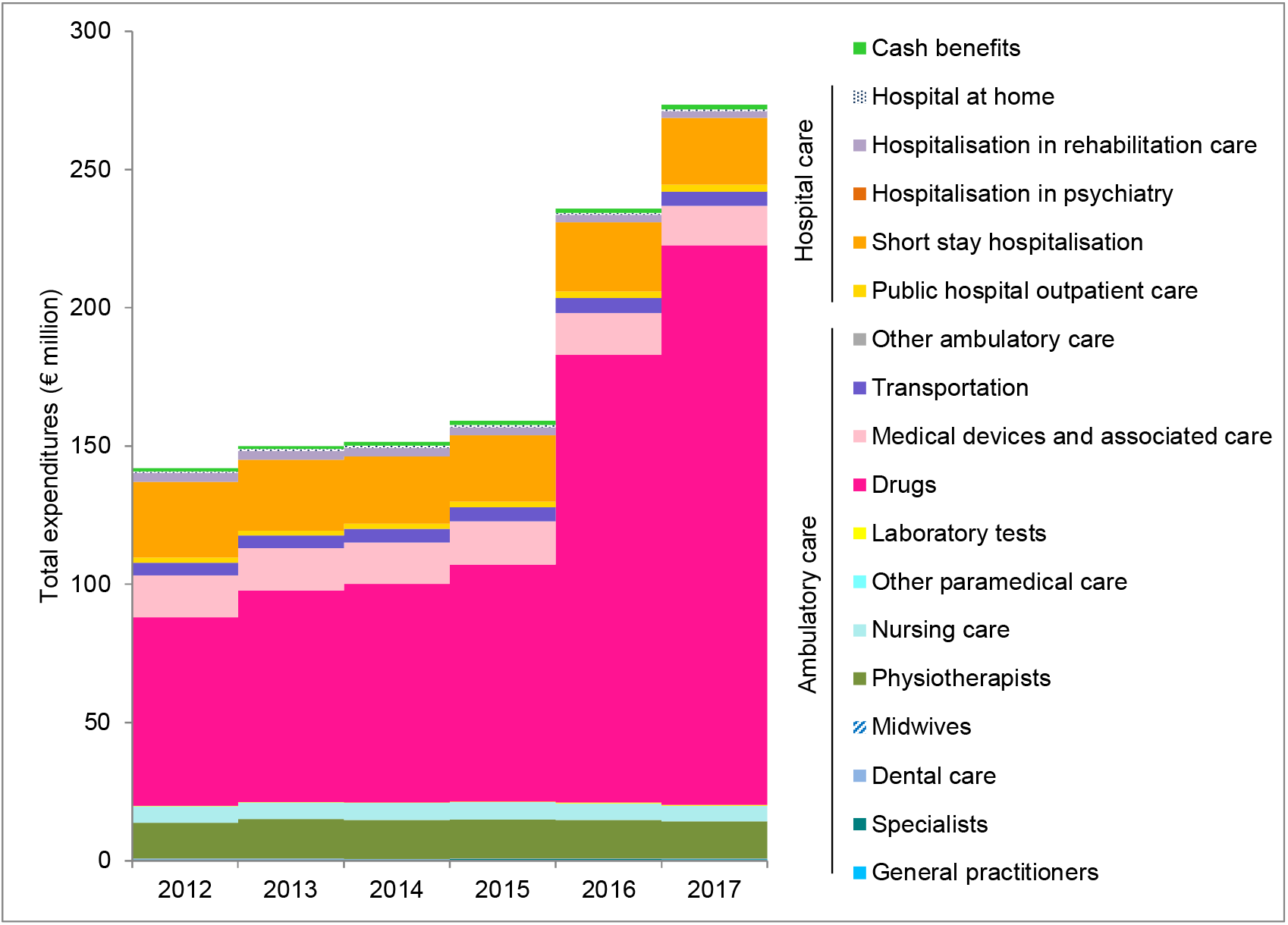
Growth of total expenditure attributed to cystic fibrosis by type of expenditure between 2012 and 2017.

